# A Consensus Transcriptional Landscape of Human End-Stage Heart Failure

**DOI:** 10.1101/2020.05.23.20110858

**Authors:** Ricardo O. Ramirez Flores, Jan D. Lanzer, Christian H. Holland, Florian Leuschner, Patrick Most, Jobst-Hendrik Schultz, Rebecca T. Levinson, Julio Saez-Rodriguez

## Abstract

2.

**Aims:** Transcriptomic studies have contributed to fundamental knowledge of myocardial remodeling in human heart failure (HF). However, the agreement on key regulated genes in HF is limited and systematic efforts to integrate evidence from multiple patient cohorts are lacking. Here we aimed to provide an unbiased consensus transcriptional signature of human end-stage HF by comprehensive comparison and analysis of publicly available datasets.

**Methods and Results:** We curated and uniformly processed 16 public transcriptomic studies of left ventricular samples from 263 healthy and 653 failing human hearts. Transfer learning approaches revealed conserved disease patterns across all studies independent of technical differences. We meta-analyzed the dysregulation of 14041 genes to extract a consensus signature of HF. Estimation of the activities of 343 transcription factors, 14 signalling pathways, and 182 micro RNAs, as well as the enrichment of 5998 biological processes confirmed the established aspects of the functional landscape of the disease and revealed novel ones. We provide all results in a free public resource https://saezlab.shinyapps.io/reheat/ to facilitate further use and interpretation of the results. We exemplify usage by deciphering fetal gene reprogramming and tracing myocardial origin of the plasma proteome biomarkers in HF patients.

**Conclusion:** We demonstrated the feasibility of combining transcriptional studies from different HF patient cohorts. This compendium provides a robust and consistent collection of molecular markers of end-stage HF that may guide the identification of novel targets with diagnostic or therapeutic relevance.

---

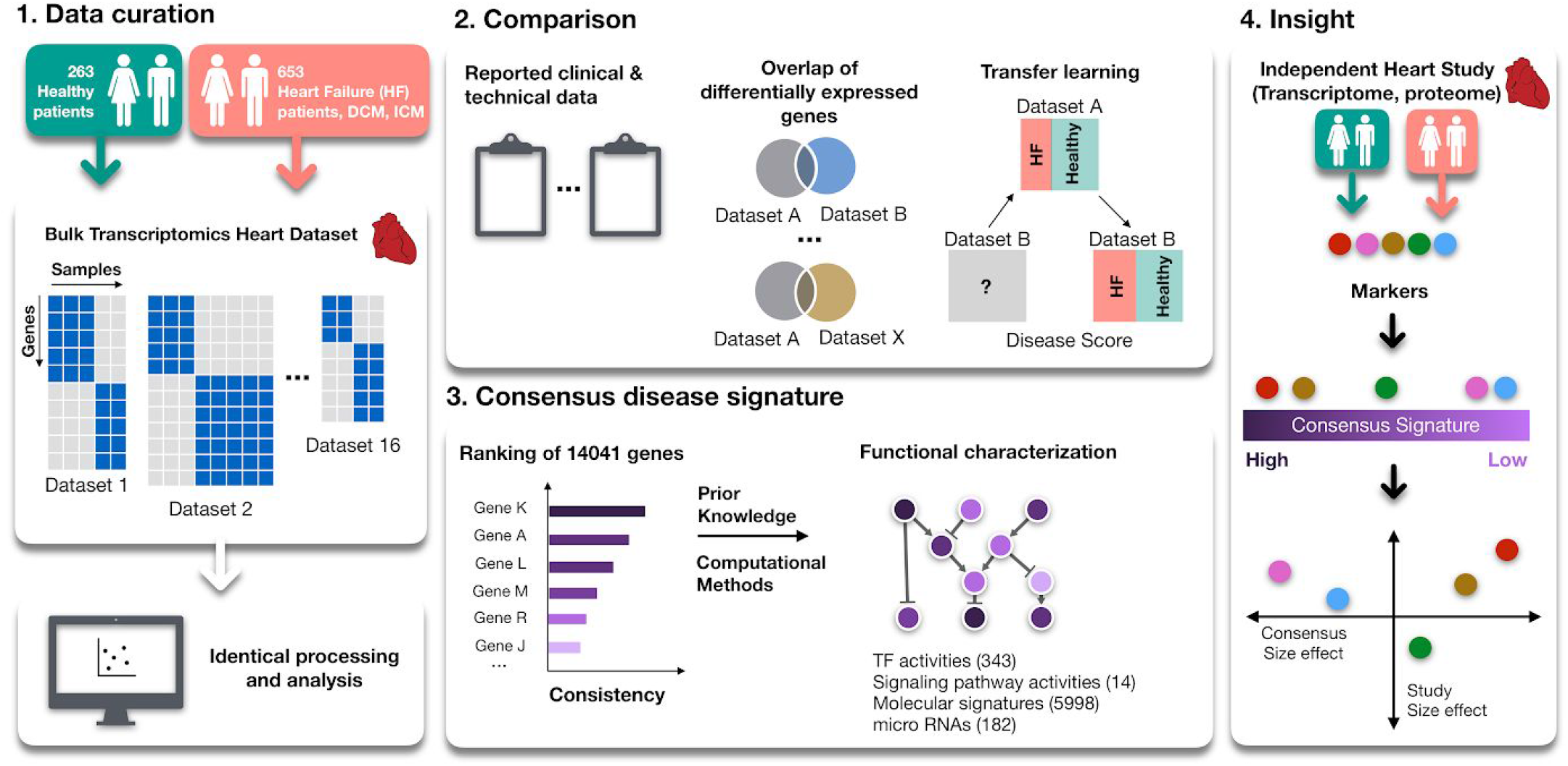

## 3. Translational Perspective

A reliable reference of the molecular processes underlying HF is needed to identify generalizable biomarkers with diagnostic or therapeutic relevance. Here, we provide a consensus transcriptional signature of human end-stage HF built from more than 900 individuals that is independent of technical biases. By tracing the myocardial origin of plasma proteomic biomarkers and defining molecular processes during the reactivation of the fetal program during HF, we demonstrate that the presented resource is crucial to complement findings in independent studies and decipher fundamental changes in failing myocardium.

## 4. Introduction

Clinical care for heart failure (HF) has not yet overcome the poor prognosis of the syndrome ^1^ To develop novel treatment and diagnostic approaches, the understanding of molecular pathophysiology of myocardial failure is crucial. Large scale transcriptomic studies have helped elucidate the complexity of gene regulation in HF, notably in processes influencing cardiac hypertrophy ^2^, reverse remodeling ^3^ and cardiac metabolism ^4^. However, low sample sizes of most studies may underestimate the effects of comorbidities, clinical history and genetic background that interact with the molecular processes active during myocardial remodeling. Lack of standards in experimental design, tissue protocols and data analysis add more confounding factors ^5^. Additionally, most transcriptomic studies lack functional interpretations, focusing in identifying a few differentially expressed genes. Therefore, the combination of multiple studies can be used to assess the robustness of patterns of gene dysregulation, identify consistent molecular changes that are less likely influenced by confounding factors, and allow for functional characterizations that can contribute to the identification of novel targets with diagnostic or therapeutic relevance.

Several reports have attempted to compare HF gene expression studies ^6-9^ but, to our knowledge, there’s no comprehensive available resource that provides a consensus transcriptional disease signature characterized with functional tools. Here, we present a meta-analysis of 16 publicly available end-stage HF transcriptome studies comprising 653 HF and 263 healthy left-ventricle biopsies. We evaluated the extent to which these geographically and technically diverse studies agree, and derived a HF consensus signature (HF-CS) that reflects robust and consistent molecular hallmarks of end-stage HF. We functionally characterized this ranking and estimated transcription factor (TF), micro RNAs (miRNAs) and signaling pathways activities that revealed established and novel insights to the transcriptional landscape of HF. Finally, we made our results publicly available to be leveraged by the research community and exemplified their utility by exploring the reactivation of fetal gene programs in HF and by tracing the myocardial origin of plasma proteomic biomarkers.

## 5. Methods

### 5.1 Study inclusion criteria

We identified human HF transcriptomic studies performed with either microarray or RNA-Seq by querying the Gene Expression Omnibus database, the European Nucleotide Archive and ArrayExpress. Search terms included “heart failure”, “ischemic cardiomyopathy”, “dilated cardiomyopathy”, “cardiac failure” and “heart disease”. We manually reviewed the results and selected studies for inclusion if: (i) case samples came from biopsies of the left ventricle of the human heart of end stage HF patients with either ischemic cardiomyopathy (ICM) or dilated cardiomyopathy (DCM); (ii) control samples were obtained from patients with non-failing hearts; (iii) data from at least 5 samples were available; (iv) microarray platforms were single channel chips and could be processed through pipelines described in section 5.2; and (v) a publication or preprint with a detailed methodology was available. The selected studies are presented in Table 1. One study (vanHeesch19) was not found by database query but literature review of cardiac gene transcription.

### 5.2 Data processing and normalization

Available raw data was downloaded to ensure consistent processing and normalization of all studies. Count matrices from RNA-Seq studies were obtained with BioJupies ^10^ and normalized with *edgeR* ^11^. Microarray studies were processed and normalized with *limma ^12^* and oligo *^13^* packages (full description in Supplemental Material).

To identify differentially expressed genes within each study, gene expression of the samples of control individuals and HF patients were compared using linear models with *limma* ^12^. Gender, age, comorbidities, etiology, occasion of sample acquisition and technical batches were used as covariates for experiments that provided this information (Supplemental Table 1). Differential expression of known markers associated with HF were used as a quality control check for all studies (Supplemental Material, SF 4).

### 5.3 Consistency between studies

We tested the degree to which individual studies could transfer their knowledge to others by defining a disease score, inspired by POE ^14^ and PROGENy ^15^. The disease score linearly combines the gene expression values of the samples of one study with the disease pattern observed in an independent reference study, captured by the t-values obtained after differential expression analysis (Supplemental Material, SF 5). Sample level disease scores were used to classify HF patients and the performance was assessed using the area under the receiver operating characteristic curve (AUROC).

### 5.4 Meta-analysis

We combined the Benjamini-Hochberg (BH) corrected p-values of the differential expression analysis for all genes that were measured in at least 10 datasets using a Fisher’s combined probability test. The degrees of freedom for the significance test of each gene were defined by the number of datasets that included it. We assumed that non-probabilistic sampling procedures happened in each study, so no additional study weighting was used ^16^. A ranking was generated based on the combined test p-values after BH correction, representing the HF-CS. The contribution of each study to the meta-analysis was estimated with the enrichment score of its top 500 differentially expressed genes in the HF-CS as calculated by gene set enrichment analysis (GSEA) ^17^.

### 5.5 Functional characterization of the HF-CS

The *-log10(meta-analysis p-value)* of each gene was weighted by its mean direction of change in all studies to create a directed HF-CS. Gene Ontology (GO) terms, canonical and hallmark pathways from MSigDB (data downloaded in December 2019) ^18^ were tested for enrichment in the directed HF-CS with GSEA ^17^ using *fgsea* ^19^ TF and miRNA activities were estimated with *viper ^20^* for human regulons obtained from DoRothEA ^21^ and the miRNA collection of targets from MSigDB ^18^, respectively. The activity of signalling pathways was calculated with PROGENy ^15^, ^22^ (Supplemental Material).

## 6. Results

### 6.1 Study curation and description

We identified 16 studies that fit the inclusion criteria (Table 1) which comprised of 263 control, 372 DCM and 281 ICM samples (Figure 1B). The studies were published between 2005 and 2019 and their sizes varied between 5 and 313 samples. Gene coverage after processing (range: 12,402 - 20,174) was comparable for all studies (mean jaccard index of ~0.67) (Supplemental Material, SF 2).

**Figure 1.**
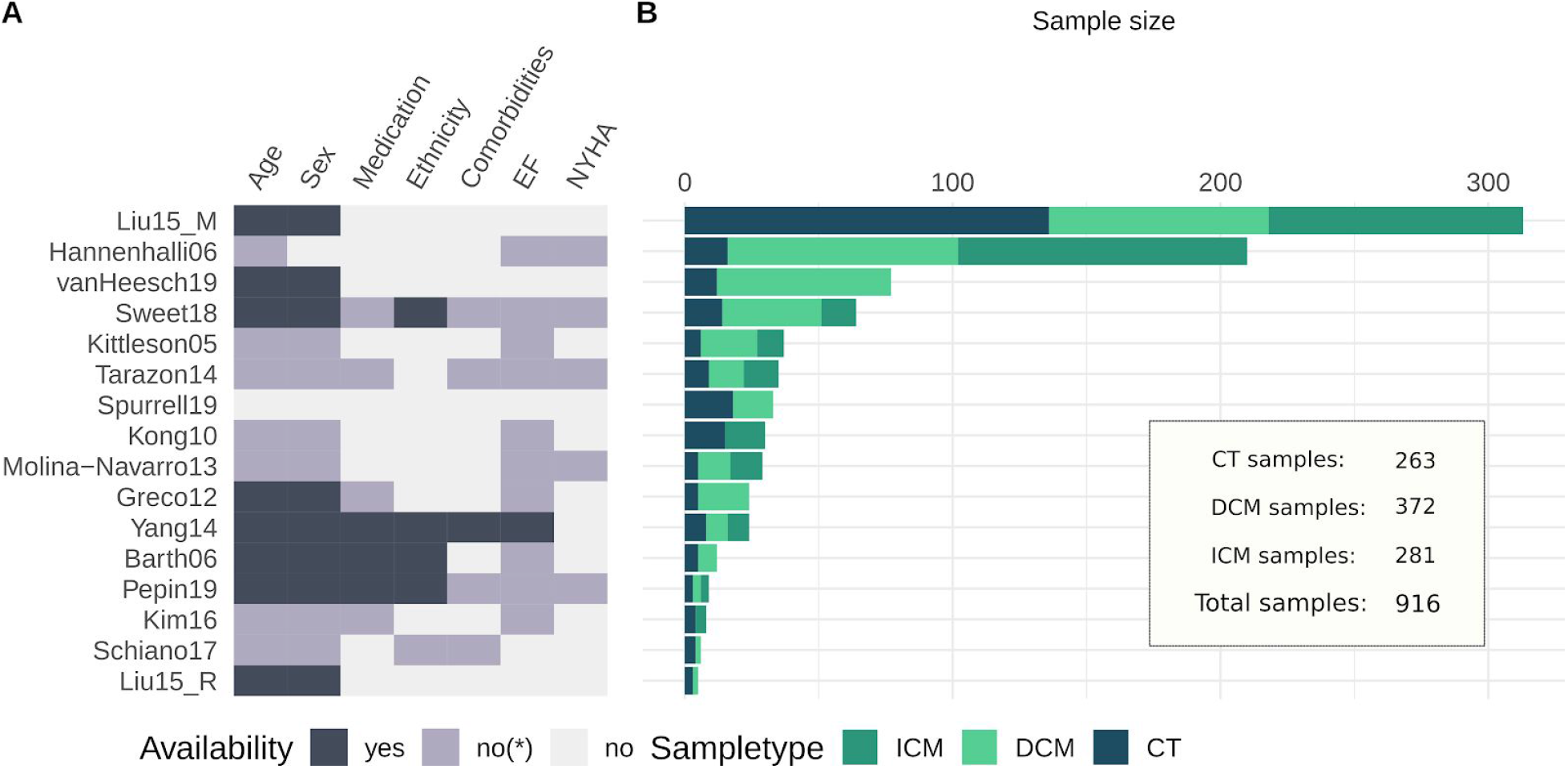
Infographic of study information. A) Sample information availability per study. yes, information per sample; no* = incomplete information or only summary statistics; no, no information available. B) Sample size comparison of studies. CT, Control.

HF samples from all studies were acquired during heart transplantation, left ventricular assist device implantation or surgical ventricular restoration which are usually performed for patients with a decompensated failing heart with reduced ejection fraction, justifying their inter-study comparability. As control samples, all studies included biopsies from donor hearts deemed unsuitable for transplant. Figure 1A displays the availability of sample information for each study concerning patient demography and HF status. When information was available, New York Heart Association (NYHA) classification ranged between III and IV and left ventricular ejection fraction (EF) was reported to be below 40% (Supplemental table 1). Age and Gender distributions are compared in Supplemental Material, SF1.

### 6.2 Sample variability and study consistency

Principal Component Analysis and Analyses of Variance were applied to various transformations of the data to evaluate sample variability across studies and etiologies (Supplemental Material). The expected interaction of technical heterogeneity with gene expression scales was observed in all samples (Supplemental Material, SF6), but was reduced by gene standardization (Supplemental Material, SF7). Only 13% of the explained variance of HF patients was captured by their etiology (Supplemental Material, SF8), suggesting a low influence at this disease stage.

We evaluated consistency across the studies by comparing their transcriptional signatures using multiple metrics. We found an almost null concordance at the gene level (mean Jaccard index of the top 500 differentially expressed genes = 0.05; Figure 3A), however, the top 500 differentially expressed genes of one study predict well HF in each other study, using sample classifications based on a disease score (median AUROC = 0.94; see Methods).

The effectiveness of the disease score to transfer knowledge of independent cohorts suggests that trends of transcriptional regulation are more stable than the expression of marker genes. To confirm this, we tested if the direction of deregulation of the top differentially expressed genes of each study were consistent with the rest. Up and downregulated genes of each study (500 in total) were enriched separately in the gene-level statistics of the collection of studies (Figure 2 C). Differentially up-regulated and down-regulated genes had a median enrichment score of 0.55 (Figure 2C, upper panel) and −0.56 (Figure 2C, lower panel), respectively. We observed a correlation between the AUROCs of the predictions and the enrichment scores of differentially expressed genes (Pearson correlations 0.48 and −0.59; p-value<10e-15, for up and downregulated genes, respectively), supporting the idea that even though the size effect of HF relevant genes are dependent on the study (Figure 2A), their direction of regulation is generally consistent, allowing their integration.

**Figure 2.**
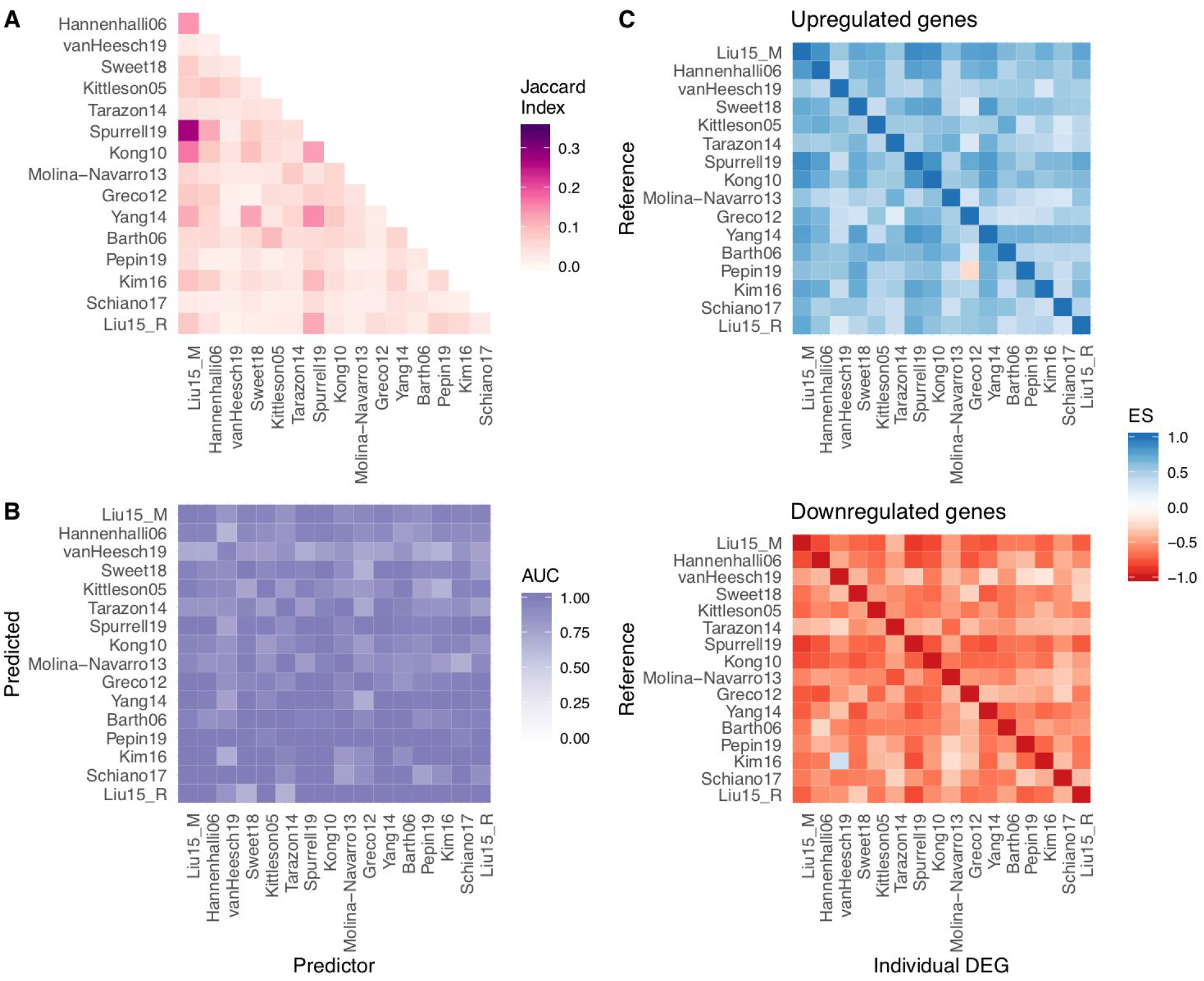
Consistency of the transcriptional signal of end-stage HF among studies. A) Pairwise-comparison of the top 500 differentially expressed genes of each study using the Jaccard index. B) AUROC of pairwise predictions using a disease score with the top 500 differentially expressed genes of each study. C) Enrichment score (ES) of the top 500 differentially expressed of each study in sorted gene level statistics lists.

### 6.3 Meta-analysis of the transcriptional responses in end-stage HF

We meta-analyzed the differential expression of 14041 genes using a Fisher combined probability test (Supplemental table 2) to create a HF-CS that captured a gradient of consistently regulated genes in end-stage HF across multiple studies regardless of their direction (Figure 3; Supplemental Material, SF10). We found no correlation between the sample size of a study and its contribution to the meta-analysis (Spearman correlation 0.24, p-value = 0.37), suggesting that proper experimental design and representative sampling could compensate for study size ^16^. Among the top 500 genes in the HF-CS (Figure 3B) we observed known HF markers such as MYH6, MME, CNN1, NPPA, KCNH2 and ATP2A2; extracellular associated proteins such as COL21A1, COL15A1, ECM2 and MXRA5; fibroblast associated protein FGF14; mast cells associated protein KIT; proteins mapped to force transmission defects like FNDC1, LAMA4, SSPN, or related to ion channels like KCNN3.

**Figure 3.**
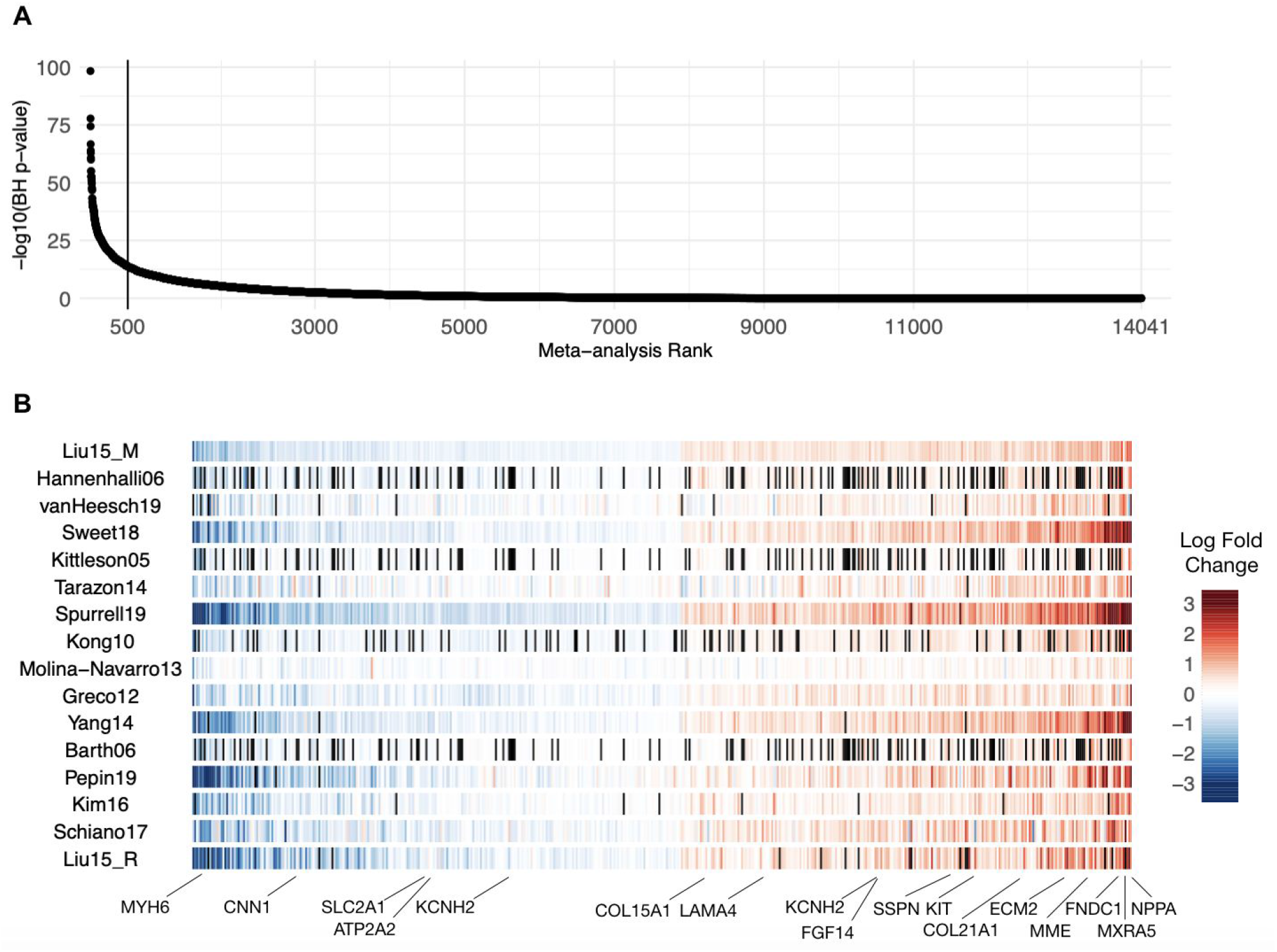
Meta-analysis summary. A) Sorted −log10(meta analysis BH p-values) of the 14041 genes included in the Fisher combined test, representing the HF-CS. B) Top 500 genes sorted by their mean log fold change across all studies, black lines represent genes that weren’t measured in specific studies. A selection of HF marker genes are highlighted.

**Figure 4.**
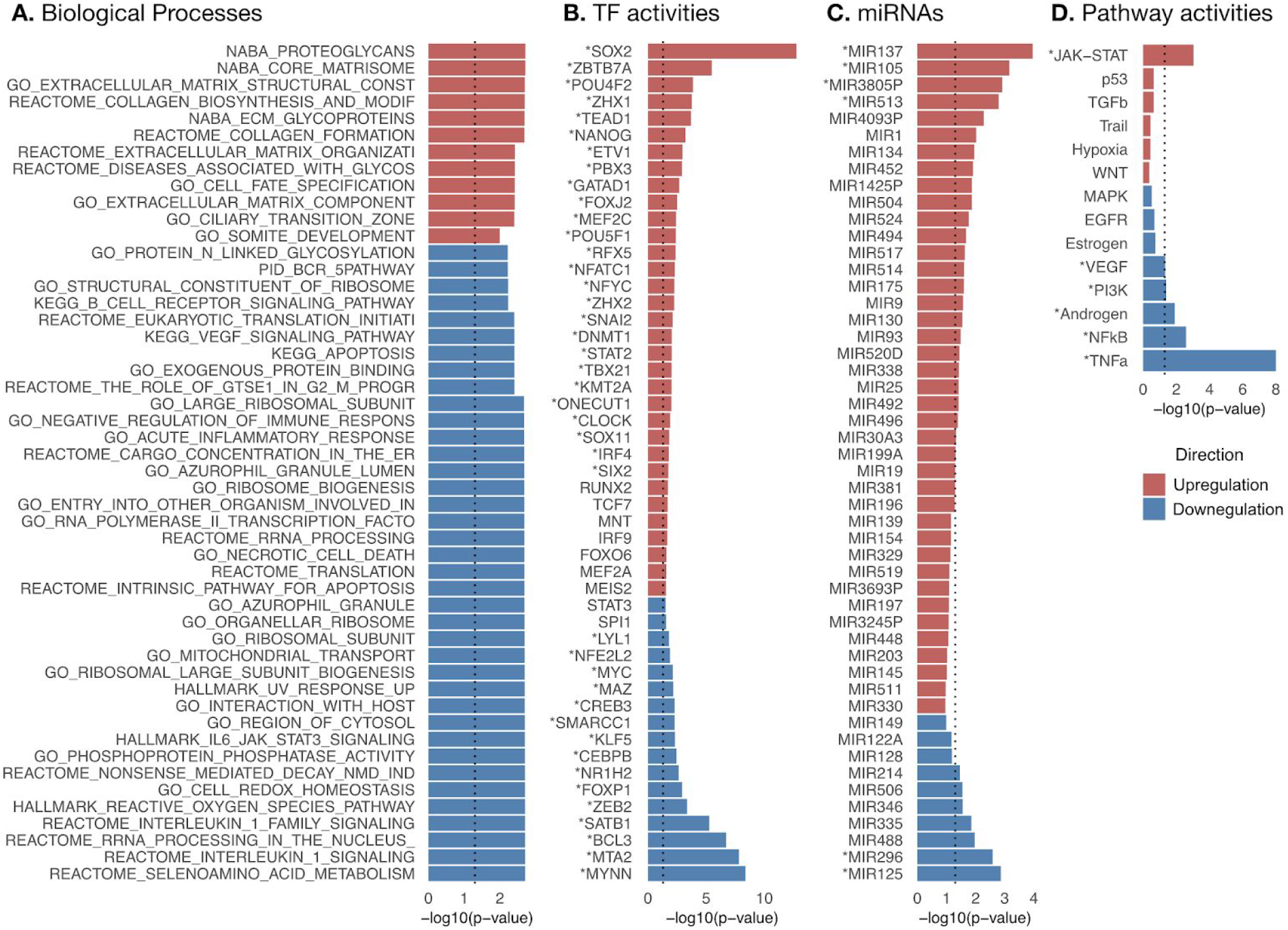
Functional characterization of the HF-CS: −log10(p-values) coloured by direction of dysregulation of the top 50 A) most enriched canonical and hallmark gene sets B) transcription factor activities, C) miRNAs’ targets, and D) all signalling pathway activities. Features with a BH-p-value<0.15 have a star next to their name. Dashed line indicates p-value = 0.05.

To evaluate the added value of the meta-analysis, we tested if the selection of the top 500 genes from the HF-CS defined a better transcriptional signature of HF than signatures obtained from individual experiments of the same size (Supplemental Material). An improvement in the AUROCs of classifiers based on the disease score was obtained (Wilcoxon paired test, p-value < 1×10e-16), and the top genes of the HF-CS were consistently more enriched in individual lists of differentially expressed genes than gene signatures from individual experiments (Wilcoxon paired test, p-value < 1×10e-16). The proportion of variability in gene expression explained by HF, controlled for other clinical and technical covariates, was greater for top genes than for genes in a lower ranking in the HF-CS (Supplemental Material; SF11).

The disease score classifier based on the HF-CS discriminated effectively HF samples of diverse etiologies (mean AUROC = 0.9) and HF samples processed with different bioinformatic pipelines (mean AUROC = 1), from studies excluded in the meta-analysis (Supplemental Table 1; Supplemental Material, SF12). These results suggest that the deregulated transcriptional processes described in our HF-CS are generalizable and may capture the disease state of diverse HF etiologies.

### 6.4 Functional evaluation of the HF-CS

We characterized the underlying deregulated processes of the HF-CS by estimating the activity of TFs, signaling pathways, and miRNAs and testing for enrichment of gene sets capturing various molecular processes (Supplemental table 3). We tested a total of 5998 gene sets, of which 579 yielded an enrichment in the HF-CS (p-value <0.05; Figure 5A). Positively enriched gene sets predominantly relate to the matrisome, while negatively enriched sets associated with diverse processes many of which involve inflammation. From the inferred transcriptional activity of 343 TFs (See Methods, Fig 5B), we found 65 TFs differentially active in HF (p-value<0.05). Among active TFs were MEF2A-C, ARNT and MEIS1-2. MEF2 family members are expressed during cardiac development and have been described to be part of the fetal reprogramming in HF ^23^. The cardiac specific depletion of ARNT resulted in an increased fatty acid oxidation leading to improved cardiac function in mice^13^. MEIS1 and MEIS2 contributed to the curbing of cardiomyocyte differentiation in mice and rats ^24,25^. Out of 211 tested miRNAs, 32 were enriched in the HF-CS (p-value <0.05) (Figure 5C), including miRNAs involved in angiogenesis (mir-126 and mir-130a^26^) and fibrosis (mir-206^27^ and mir-214^28^). From the estimated signaling pathway activities (Figure 5D), TNFα, NFKB, Androgen receptor, PI3K and VEGF signaling were consistently inactive (p-value<0.05). While TNFα levels are elevated in HF patients in relation to decreasing functional status^29^, clinical trials targeting TNFα failed to improve HF outcome ^30^. Additionally, there is recent evidence that TNFα signaling could be part of a physiological inflammatory response exerting cardioprotective effects ^31,32^. Down regulated pathway activities of TNFα and NFKB accompanied by decreased TF activities of RELA and NFKB1 in the HF-CS indicate an ambiguous role of TNFα during HF that has not been fully appreciated yet. JAK-STAT was the only pathway with a high activity (p-value<0.05). The JAK-STAT pathway is activated by growth factors and cytokines and is an imperative regulator of cardiac development and inflammation. The role of JAK-STAT in HF is ambivalently discussed ^33^, with evidence that JAK-STAT is involved in cardiac hypertrophy ^34^, ischemic pre and post conditioning ^35^ and cardiac fibrosis ^36^. Taken together, the functional interpretation of the HF-CS reflects molecular and cell biology perturbations that shape the pathological gene expression profile in HF and therefore reveals promising objects of future investigations.

**Figure 5.**
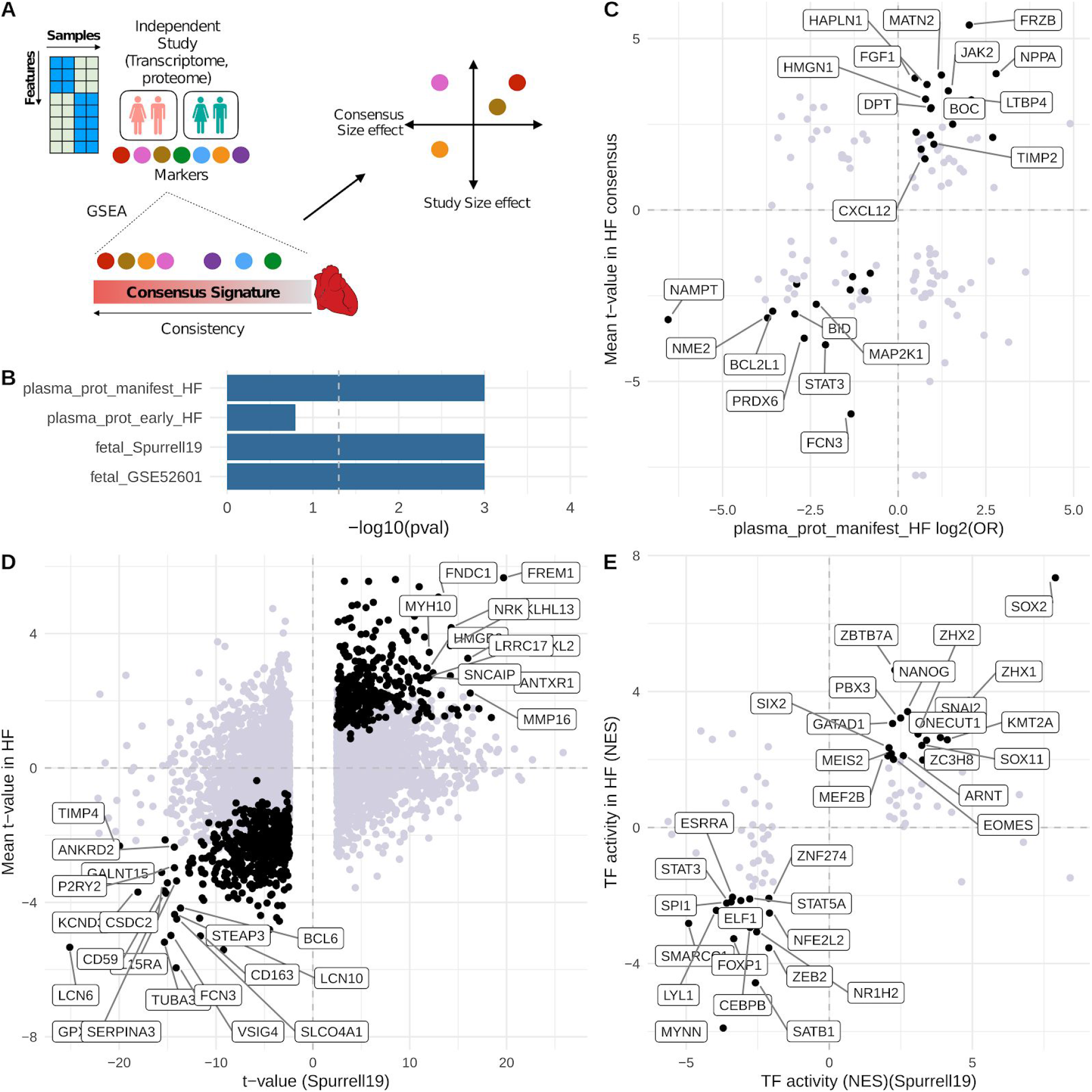
HF-CS as a reference that complements independent studies: A) Schematic of a suggested framework. Marker features from independent studies are enriched in the HF-CS with GSEA. Features that belong to the leading edge are further filtered, e.g. by correlation or ranking in the HF-CS. B) Enrichment results of marker features from four individual studies. C) Plasma proteome of HF patients mapped to the HF-CS. D) Fetal cardiac transcriptome (Spurrell19) mapped to HF-CS on gene level and E) TF level. Black dots in C & D indicate correlated features in the enrichment leading edge; labeled features in C & D indicate genes with a rank < 500 in HF-CS. Black dots in E indicate overlap with significantly dysregulated TFs derived from the HF-CS.

### 6.5 HF-CS as a resource for biomarker detection and hypothesis building

Finally, we tested how the HF-CS could be leveraged to build or confirm hypotheses from independent studies by comparing the dysregulation patterns of their reported markers (Figure 5A, Supplemental Material, Supplemental table 4). We analyzed the plasma proteome of early and manifest HF patients from Egerstedt, *et al^37^* to trace their myocardial origin. We observed a clear enrichment of manifest HF proteins (GSEA p-value = 0.0001) and a modest enrichment of early HF proteins (GSEA p-value = 0.13) in the top of the HF-CS (Figure 5B). 64 plasma proteins from manifest HF were part of the enrichment leading edge and agreed with the direction of transcriptional regulation (Figure 5C), including the established HF marker NPPA and novel potential markers like extracellular matrix components (HAPLN1, MATN2, COL8A1), Wnt signaling antagonists (SFRP1, FRZB) and cytokines (CXCL12). Additionally, we dissected the reactivation of fetal gene programs in HF by analyzing two public fetal cardiac transcriptomes (GSE52601, Spurrell19) and their estimated TF activities. Fetal transcriptional signatures of both studies were enriched in the top rankings of the HF-CS (GSEA p-value < 0.01) (Figure 5B). 221 of the top 500 genes from HF-CS correlated with fetal genes reported by Spurrell19 (Figure 5D) while 32 TFs correlated between fetus heart and HF-CS (Figure 5E). Similar results were observed for GSE52601 (Supplemental Material, SF13).

## 7. Discussion

In this study we present a comprehensive meta-analysis of the HF transcriptome, comparing 16 datasets and a total of 916 samples. HF is a complex disorder both on the clinical and genetic levels. As such, the published work in myocardial transcriptomics represents a heterogeneous picture of transcriptional regulation in the heart. In the studies included in this meta-analysis, clinical heterogeneity is compounded by wide variability in analysis pipeline, study design, tissue protocol, and patient selection. Our work shows that despite these difficulties, combining these studies provides an opportunity not only to robustly evaluate their reproducibility, but also to gain a more complete picture of transcriptional regulation. To our knowledge, this report represents the largest comprehensive meta-analysis of human HF transcriptome studies to date.

One strength of this study is the added robustness to the gene dysregulation associations found in end-stage HF, based on integrating equally the evidence of a diverse collection of studies. In this meta-analysis we balanced the bias of the experimental design, and increased the sample size, while reducing technical variance by standardizing the bioinformatics processing and analysis of each dataset. Another strength is that the estimation of TF and signaling activities provides a functional catalog of interpretable features that are more robust than individual markers, since they integrate the expression pattern of multiple genes.

Important limitations of our study relate to the data used. In this meta-analysis, we only included public datasets that may misrepresent the population of HF patients. A second limitation is the lack of reported demographic and clinical data. As the necessity of studying HF in clinically ramified subgroups is becoming evident ^38^, the impact of comorbidities, medication and disease phenotype on the gene transcription profile needs to be considered. Despite these limitations, we believe that the sample size in our meta-analysis and the application of novel functional transcriptomic tools can provide useful insights into the pathophysiology of HF. Furthermore, our methodology is general and, as more data becomes available, the meta-analysis can be expanded.

The presented HF-CS describes established hallmarks in HF, including fetal reprogramming, cardiac fibrosis, activation of JAK-STAT and depletion of VEGF signaling. This encourages us to highlight findings that have been less explored yet, like the role of active TFs including MEIS1-2, ARNT, RUNX2, TEAD1 or the absence of TNFα signaling.

We built the user-friendly free platform “ReHeaT - Reference of the Heart failure Transcriptome” (https://saezlab.shinyapps.io/reheat/) with all results to facilitate further interpretations of the HF-CS and to provide a trustworthy resource for biomarker detection and hypothesis building or confirmation. As transcriptomic technologies move towards single cell and spatial resolution, this resource could help to confirm cell-type specific elements in a large HF population.

We demonstrated the utility of the meta-analysis by integration with studies analyzing the fetal transcriptome and the plasma proteome from HF patients. The activation of a fetal gene program has been linked to the molecular remodelling processes in HF. However, detailed pathophysiology of this process is incompletely understood. Our analysis provides a plethora of genes and TFs that might shape the fetal response in HF. We detected established TFs like MEF2, but also identified a collection of less explored TFs including SOX2, ZBTB7A, NANOG, ONECUT1. The plasma proteome of HF patients is used to identify circulating biomarkers. However, tracing the origin of measured candidates to the heart is often difficult. We filtered circulating proteins based on the HF-CS and identified the established marker NPPA^39^. Other identified markers include Wnt modulators SFRP1 and FRZB, the latter has been associated with heart failure outcome before ^40^. We also suggest CXCL12 plasma levels to be of myocardial origin, which is associated with stroke ^41^ and acute heart failure ^42^. HAPLN1, MATN2 and COL8A1 constitute extracellular matrix components with to date an unknown role in HF. These candidates could represent biomarkers of pathophysiological relevance and potential clinical utility.

In summary, we demonstrated the feasibility of combining gene expression data sets from different technologies, years and centers in a biological meaningful way. We highlight the importance of data sharing by building a rich resource and displaying its utility to advance HF research. As the number of cardiovascular high-throughput studies increases, the need of structured data integration is evident. We provide a reference for this purpose that is applicable to many other research topics within the cardiovascular field.

## Data Availability

Processed data can be downloaded from Zenodo at https://zenodo.org/record/3797044#.XsQPMy2B2u5.

## 8. Acknowledgments

We thank Tim Kuhn, Martin Busch, and Jakob Wirbel for useful discussions. We thank Hyojin Kim for editing the graphical abstract.

## 11. Appendices

### 12. Text tables

**Table 1.** Overview of studies selected for meta analysis. 16 data sets fulfilled the inclusion criteria. Samples size is displayed after processing. GEO, gene expression omnibus; CT, control; HF, heart failure.

| Study ID | GEO ID | Samples<br>(CT + HF) | Technology | Year | Country | Citation |
| --- | --- | --- | --- | --- | --- | --- |
| Liu15_M | GSE57345 | 313 | Microarray | 2015 | USA | <sup>43</sup> |
| Hannenhalli06 | GSE5406 | 210 | Microarray | 2006 | USA | <sup>44</sup> |
| vanHeesch19 | not in GEO | 77 | RNAseq | 2019 | Germany | <sup>45</sup> |
| Sweet18 | GSE116250 | 64 | RNAseq | 2018 | USA | <sup>46</sup> |
| Kittleson05 | GSE1869 | 37 | Microarray | 2005 | USA | <sup>47</sup> |
| Tarazon14 | GSE55296 | 35 | RNAseq | 2014 | Spain | <sup>48</sup> |
| Spurrell19 | GSE126573 | 33 | RNAseq | 2019 | USA | <sup>49</sup> |
| Kong10 | GSE16499 | 30 | Microarray | 2010 | USA | <sup>50</sup> |
| Molina-Navarro13 | GSE42955 | 29 | Microarray | 2013 | Spain | <sup>51</sup> |
| Greco12 | GSE26887 | 24 | Microarray | 2012 | Italy | <sup>52</sup> |
| Yang14 | GSE46224 | 24 | RNAseq | 2014 | USA | <sup>53</sup> |
| Barth06 | GSE3585 | 12 | Microarray | 2006 | Germany | <sup>54</sup> |
| Pepin19 | GSE123976 | 9 | RNAseq | 2019 | USA | <sup>55</sup> |
| Kim16 | GSE76701 | 8 | Microarray | 2016 | USA | <sup>56</sup> |
| Schiano17 | GSE71613 | 6 | RNAseq | 2017 | Italy | <sup>57</sup> |
| Liu15_R | GSE57344 | 5 | RNAseq | 2015 | USA | <sup>43</sup> |

### 13. Figure

### 14. Supplementary files

Submitted in separate files:

1. **Supplemental table 1**. Complete description of the studies included in the meta-analysis
2. **Supplemental table 2**. Summary statistics and rankings from the meta-analysis.
3. **Supplemental table 3**. Functional characterization of the consensus signature. GSEA gene set level statistics for MSigDB’s canonical pathways and gene ontology terms, DoRothEA’s transcription factor level statistics, PROGENy’s signalling pathway level statistics, and micro-RNA level statistics.
4. **Supplemental table 4**. Full results from validation analysis.

### 15. Data and code availability

Code used for all analyses is available at https://github.com/saezlab/HFmeta-analysis/ and processed data can be downloaded from Zenodo at https://zenodo.org/record/3797044#.XsQPMy2B2u5

